# Detection of Bourbon virus specific serum neutralizing antibodies in human serum in Missouri, USA

**DOI:** 10.1101/2022.03.17.22272570

**Authors:** Gayan Bamunuarachchi, Houda Harastani, Paul W. Rothlauf, Ya-nan Dai, Ellebedy Ali, Daved Fremont, Sean P.J. Whelan, David Wang, Adrianus C. M. Boon

**Author notes:** Equal contribution (Gayan Bamunuarachchi, Houda Harastani, Paul W. Rothlauf). Lead Contact Adrianus C. M. Boon, Department of Internal Medicine, Washington University in Saint Louis, School of Medicine, St. Louis, MO 63110.

## Abstract

Bourbon virus (BRBV) was first discovered in 2014 in a fatal human case. Since then it has been detected in the *Amblyomma americanum* tick in the states of Missouri and Kansas in the United States. Despite the high prevalence of BRBV in ticks in these states, very few human cases have been reported, and the true infection burden of BRBV in the community is unknown. Here, we developed two virus neutralization assays, a VSV-BRBV pseudotyped rapid assay, and a BRBV focus reduction neutralization assay, to assess the seroprevalence of BRBV neutralizing antibodies in human sera collected in 2020 in St. Louis, Missouri. Out of 440 human serum samples tested, three (0.7%) were able to potently neutralize both VSV-BRBV and wild type BRBV. These findings suggest that human infections with BRBV are more common than previously recognized.

**Importance:** Since the discovery of the Bourbon virus (BRBV) in 2014, a total of five human cases have been identified, including two fatal cases. BRBV is thought to be transmitted by the Lone Star tick, which is prevalent in the East, Southeast, and Midwest of the United States (US). BRBV has been detected in ticks in Missouri and Kansas and serological evidence suggests that it is also present in North Carolina. However, the true infection burden of BRBV in humans is not known. In the present study, we developed two virus neutralization assays to assess the seroprevalence of BRBV specific antibodies in human sera collected in 2020 in St. Louis, Missouri. We found that a small subset of individuals is seropositive for neutralizing antibodies against BRBV. Our data suggest that BRBV infection in humans is more common than previously thought.

## INTRODUCTION

Emerging and reemerging infectious diseases cause substantial global health and socio-economic burden, and have a significant impact on human and animal life. Viral pathogens, like Ebola virus, severe acute respiratory syndrome coronavirus-2 (SARS-CoV-2), influenza virus, and Chikungunya virus, are among the most prominent emerging zoonotic infections [1]. Many zoonotic diseases are transmitted by arthropod vectors such as mosquitos and ticks and are an important cause of human morbidity and mortality in the United States (US). Ticks are the main vector that transmit pathogens in the US including four different viruses; Powassan virus, Colorado tick fever virus, Heartland virus (HRTV), and Bourbon virus (BRBV).

BRBV belongs to the family *Orthomyxoviridae*, genus *Thogotovirus*. BRBV is a negative-sense segmented RNA virus whose genome is composed of 6 gene segments [2]. Segment 4 encodes the viral envelope glycoprotein (GP) that is necessary for virus attachment and entry into cells [3], and it is the main target for virus neutralizing antibodies [4]. A total of five human BRBV cases, including two fatal cases, have been reported since the discovery of this virus in 2014. The first case of BRBV was an adult male patient from Bourbon County, Kansas, USA. The patient was hospitalized with febrile illness and later died from renal failure and acute respiratory distress syndrome. Subsequent culturing and next-generation sequencing of the blood of this patient identified BRBV [5]. In 2017, a state park official from Missouri was diagnosed positive for BRBV. This patient later died of respiratory failure and cardiac complications [2, 6]. Both patients had reported tick exposure and bites prior to becoming ill [2, 5].

BRBV is isolated from lone star ticks (*Amblyomma americanum)* [7-10]. These ticks are abundant and aggressive human-biting ticks that are widely distributed across the East, Southeast, and Midwest US [8, 11]. Rabbits fed on by BRBV-infected ticks developed high antibody titers to the virus suggesting the lone star tick is a competent vector of BRBV [10]. BRBV neutralizing antibodies have been identified in different wild animal species. Using plaque-reduction neutralization test (PRNT), 50% and 86% seroprevalence of BRBV neutralizing antibodies was found in sera from raccoons and white-tailed deer in Missouri respectively [12]. Moreover, 56% seroprevalence was observed in white-tailed deer sera from North Carolina [13]. These observations demonstrate that the rate of infection of wild animals is significant, raising questions as to the true infection rate and clinical burden of BRBV in humans.

Serology can provide an important benchmark on population immunity against pathogens. However, to date there have been no serosurveillance studies assessing the seroprevalence of BRBV specific neutralizing antibodies in humans in the US and world. Thus the objectives of our study were to develop BRBV neutralization assays and measure the human seroprevalence of BRBV infection. We found that nearly 1% of the human sera, obtained in 2020, contained BRBV neutralizing antibodies. These data suggest that BRBV infection in people is more common than previously thought.

## MATERIALS AND METHODS

### Virus and cell culture

Vero E6 and Vero-CCL81 cells were maintained in Dulbecco’s Modified Eagle Medium (DMEM) containing 4.5 g/L glucose, L-glutamine, and sodium pyruvate (Corning) and supplemented with 10% Fetal Bovine Serum (FBS). Vero E6 cells (ATCC) were grown in DMEM media (Corning Cellgro) supplemented with 5% FBS (Biowest), 100 U/mL Penicillin (Life Technologies), 100 μg/mL streptomycin (Life Technologies), and 2 mM L-Glutamine (Corning). Expi293F cells (Thermo Fisher Scientific) used for transfection were maintained in Expi293 Expression Medium (Gibco).

VSV-eGFP-BRBV (herein VSV-BRBV) was generated by replacing the endogenous G gene in a molecular cDNA of vesicular stomatitis virus (VSV), encoding eGFP, with a codon-optimized version of the BRBV GP gene (Original strain, Kansas, Genbank AMN92169.1). Virus rescue was performed as described [14]. Briefly, BSRT7/5 cells were infected with vaccinia virus vTF7-3 and subsequently transfected with T7-driven helper plasmids encoding VSV N, P, L, and G, and the VSV-BRBV cDNA. Cell supernatants were harvested after 72 hours, centrifuged at 1,000 x g for 5 min, and passed through a 0.22 μm filter. Vero-CCL81 cells were then infected with rescue supernatants, and plaques in agarose plugs were isolated and passed to Vero-CCL81 cells for p1 stock generation. Working stocks were obtained by growing VSV-BRBV on Vero-CCL81 cells at a multiplicity of infection of 0.01. RNA was extracted from virus stocks using the QIAamp Viral RNA Mini Kit. RNA was reverse-transcribed using the QIAGEN OneStep RT-PCR Kit with primers targeting VSV M-G (forward) and G-L (reverse) intergenic regions. Stock sequences were confirmed by Sanger sequencing. BRBV (strain BRBV-STL) [2] was grown on Vero E6 cells. Virus stocks were sequenced by next generation, aliquoted, stored at -80°C, and used for all subsequent studies.

### VSV-BRBV characterization

Vero-CCL81 cells were infected with eGFP-expressing VSV (herein referred to as VSV) or VSV-BRBV at an MOI of 1. Cells were imaged 6 hours post infection (hpi). Plaque assays were performed by infecting Vero-CCL81 cells with the aforementioned viruses for 1 h at 37°C, after which the inoculum was removed, and cells were overlayed with an agarose-containing overlay. Plates were imaged 24 hpi using a biomolecular imager in the FITC channel.

### Recombinant proteins

Recombinant BRBV GP (rGP, residues 20-485) was expressed using the baculovirus expression system or transiently expression in mammalian cell lines. For expressing with baculovirus expression system, the gene block of BRBV GP ectodomain followed by a trimerization sequence (GYIPEAPRDGQAYVRKDGEWVLLSTFL) from the bacteriophage T4 fibritin and 6*His tag at the extreme C-terminal was cloned into a modified pOET1 (Mirus Bio) baculovirus transfer vector containing green fluorescent protein as an indicator [15]. Transfection and amplification were carried out according to the *flash*BAC baculovirus expression system manual (Mirus Bio). The High Five™ (Gibco) cell culture supernatant was harvested after 72 hpi, and concentrated before dialysis against buffer 20 mM NaHCO3, 300 mM NaCl, pH 8.0. BRBV GP ectodomain was captured by passaging the supernatant over Ni^2+^ affinity resin (GoldBio) and eluted with 500 mM imidazole (Sigma-Aldrich). The recovered proteins were further applied to HiLoad 16/600 Superdex 200 column (GE healthcare) in 20 mM HEPES, 150 mM NaCl, and pH of 7.5. For expression in mammalian cells, the gene fragment harboring the ectodomain, the trimerization domain as well as 6x-His tag from the recombinant baculovirus vector was cloned into pFM1.2R vector [16] and transiently expressed in Expi293F cells. The cell culture supernatant was harvested after 5 days post-transfection and concentrated before dialyzing against buffer 20 mM Tris-Cl pH 8.5, 150 mM NaCl. Then the BRBV GP ectodomain protein was purified with the same strategy as described above, with a combination of Ni affinity chromatography and size exclusion chromatography.

### Animal experiments

All animal experiments were preformed according to the Institutional Animal Care and Use Committee (IACUC) at Washington University in St. Louis. Eleven-to-twelve weeks-old female C57BL/6 mice were obtained from Jackson Laboratories. Mice (n *=* 3) were immunized intramuscularly with 500 ng of Beta-propiolactone (BPL; Sigma-Aldrich) inactivated and purified BRBV complemented with addavax (InvivoGen) (1:1, *v:v*) to generate BRBV GP specific monoclonal antibodies (mAb). Four weeks later, the mice were immunized intramuscularly with 5 µg of baculovirus-derived recombinant GP of BRBV complemented with addavax (1:1, *v:v*). Sera was collected one day prior to each immunization and stored at -20°C. To generate positive control sera, containing BRBV-specific neutralizing antibodies, we infected C57BL/6 mice with 10^4^ infectious units of BRBV and collected sera at 22 days post infection. Sera from five BRBV-infected mice were pooled together to use as the positive control in the virus neutralization assays.

### Flow cytometry

For B cell staining, spleen cells were collected five days post final immunization and stained for 30 min on ice with CD16/CD32 (93; Biolegend^®^ 1:100) and biotinylated mammalian expressed rGP (1:100) diluted in PBS supplemented with 2% FBS and 2 mM EDTA (P2). Next, the cells were washed twice with P2 and stained for 30 min on ice with anti-CD19-FITC (1D3; BioLegend^®^ 1:100), anti-IgD-APC-Cy7-A (11-26c.2a; BioLegend^®^ 1:100), anti-Fas-PE-A (Jo2; BD Biosciences 1:200), anti-GL7-PerCP-Cy5.5-A (GL7; BioLegend^®^ 1:50), anti-CD71-PE-Cy7-A (RI7217; BioLegend^®^ 1:100), anti-CD138-BV421 (281-2; BioLegend^®^ 1:100), anti-CD4-AF700 (GK1.5; BioLegend^®^ 1:100) and Zombie Aqua (BioLegend^®^ 1:200) diluted in Brilliant Stain buffer (BD Horizon). After 30 min, the cells were washed twice with P2, after which they were analyzed and single rGP-binding antibody-secreting B-cells (ASC; CD4^-^, CD19^+^, IgD^lo^, CD95^+^, GL7^-^, CD138^+^, BRBV-GP^+^ live singlet lymphocytes) were sorted (n = 376) into four 96-well plates containing 2 μL of Lysis Buffer (Clontech) supplemented with 1 U/μL RNase inhibitor (New England BioLabs) with a FACSAria II and immediately frozen on dry ice. Flow cytometry data were analyzed using FlowJo™ v.10.8.1.

### Monoclonal antibody production

mAbs were generated as previously described previously [17]. Briefly, variable heavy (VH) and variable light kappa and lambda (Vκ and Vλ, respectively) genes were amplified by RT-PCR and nested PCR from single-cell sorted BRBV-GP specific ASCs using mixtures of primer sets specific for IgG, IgM/A, Igκ, and Igλ using first-round and nested primer sets PCR products were then loaded on a 1% agarose gel (Lonza), purified and then sequenced. Sequencing data was annotated using IMGT/V-QUEST tool v3.5.28 on the ImMunoGenetTics database (available at http://www.imgt.org/IMGT_vquest/) [18, 19]. Clonally related cells were identified by similarity in variable heavy and light chain usage, CDR3 length, and amino-acid composition. To generate recombinant mAbs, heavy chain V-D-J and light chain V-J fragments were PCR amplified from first-round PCR products with mouse variable gene forward primers and joining J gene reverse primers having 59 extensions for cloning by Gibson assembly as previously described [20] and were cloned into pABVec6W Ab expression vectors [21] in frame with either human IgG1, Igκ, or Igλ constant region. Expression plasmids were sequence verified and transfected at a 1:2 ratio of heavy to light chain ratio into Expi293F cells using the Expifectamine 293 Expression Kit (Thermo Fisher Scientific). Culture supernatant was collected seven days post transfection and mAbs, secreted into the supernatant, were purified with protein A agarose (Invitrogen), and then stored at 4°C until further used.

### Enzyme-linked immunosorbent assay (ELISA)

Ninety-six well microtiter plates (Thermo Fisher Scientific) were coated overnight at 4°C with 1 µg/mL of baculovirus expressed rGP in PBS. After washing the wells 3x with 280 µL PBS-T (PBS supplemented with 0.05% Tween-20), wells were blocked with 280 µL of PBS supplemented with 10% FBS and 0.05% Tween-20 (blocking buffer) for 1.5 h at room temperature (RT). Blocking buffer was removed, and 3-fold serial dilutions starting from 1:30 for mouse polyclonal sera or 30 µg/mL for mAbs were added and incubated for 1 h at RT. An influenza A virus specific mAb (1C11) specific for the hemagglutinin of A/Puerto Rico/8/1934 was used as a negative control. Plates were washed 3x with PBS-T. mAbs were detected using goat-anti-human IgG-HRP (Jackson ImmunoResearch, Cat no. 109-035-088) while mouse sera were detected using goat-anti-mouse IgG1-HRP (Southern Biotech, Cat no. 1070-05) diluted 1:1000 in blocking buffer for 1 h at RT. After washing 3x with PBS-T followed by 3x with PBS, the assay was developed with 100 µL of substrate solution (phosphate-citrate buffer with 0.1% H2O2 and 0.4 mg/mL o-Phenylenediamine dihydrochloride (OPD, Sigma-Aldrich)) was added to all wells and incubated for 5 min before the reaction was stopped with 1M HCl (100 µL). Optical density measurements were taken at 490 nm using a microtiter plate reader (BioTek).

### Focus forming assay (FFA)

Monolayers of Vero E6 cells, seeded in 96 well tissue culture treated plates, were washed once with PBS and incubated with VSV-BRBV or BRBV diluted in serum free DMEM (Corning). After 1 h at 37°C, the inoculum was aspirated, the cells were washed with PBS, and 100 µL of 1% methylcellulose (Sigma) in 1x minimum essential media (MEM, Corning) supplemented with 2% FBS (Biowest) was added to each well. The cells were incubated for 24 h at 37°C/5% CO2 before the monolayer was fixed with 100 μL of 5% Formalin (Fisher Chemicals) for 1 h at RT. Cells were washed with PBS and subsequently incubated with the BRBV-GP specific mAb E02 in PBS-T plus 1% bovine serum albumin (BSA, Sigma). After 1 h incubation at RT, the cells were washed three times with PBS-T and incubated with HRP-conjugated goat anti-human IgG (Sigma) diluted in PBS-T + 1% BSA for 1 h at RT. Finally, cells were washed three times with PBS-T, and VSV-BRBV or BRBV infected cells were visualized using TMB substrate (Vector laboratories) and quantitated on an ImmunoSpot^®^ Analyzer (Cellular Technologies).

### Human serum cohort

We screened 440 adult serum samples that were part of a set of residual samples sent to Barnes-Jewish Hospital, Missouri for physician-ordered vitamin D testing between 27 April 2020 and 12 May 2020 [22]. This study was approved by the Human Research Protection Office at Washington University in St. Louis (approval no. 202004199).

### Rapid eGFP-based VSV-BRBV-GP virus neutralization assay

Vero E6 cells were cultured overnight in tissue culture treated black 96-well plates (Corning). Human sera were heat-inactivated at 56°C for 30 min and serially diluted 1:10 and 1:30 in serum free DMEM (DMEM-SF). The next day 200 focus forming units (FFU) of VSV-BRBV virus diluted in an equal volume of DMEM-SF were added to the diluted human serum samples (final serum dilution is 1:20 and 1:60) and incubated for 1 h at RT. Next, the antibody virus complexes were added to the Vero E6 cells that were washed once with PBS and incubated for 8 h at 37°C. Each serum sample and dilution was tested in duplicate and each assay included a positive control mouse serum sample, and no serum controls (DMEM media only). After 8 h the cells were fixed with an equal volume of 10% formalin containing 10 µg/mL Hoechst (Sigma) was added to each well for 1 h at RT. Subsequently, cells were washed once with PBS, and images were acquired with the InCell 2000 Analyzer (GE Healthcare). This automated microscope contains both FITC and DAPI channels to visualize infected cells (i.e. eGFP-positive cells) and nuclei respectively. Each well is divided into 4 fields that are imaged with a 4x objective lens. Subsequently, cumulative eGFP-positive cells and nuclei of the 4 fields were counted. Images were analyzed using the Multi-Target Analysis Module of the InCell Analyzer 1000 Workstation Software (GE Healthcare). eGFP-reduction neutralization activity of the serum samples was calculated by dividing the number of eGFP positive cells for each serum sample by the average number of eGFP positive cells in the no serum control wells.

### BRBV neutralization assay

A focus reduction neutralization assay (FRNT) was developed for BRBV. Vero E6 cells were seeded overnight in tissue-cultured treated 96-well plates (Corning). Heat-inactivated human or mouse sera were diluted 1:20 in DMEM-SF media and subsequently serially diluted 3-fold to a final dilution of 1:14580. Next, an equal volume of DMEM-SF containing 200 FFU of BRBV was added to each serum dilution and incubated for 1 h at RT. The addition of the virus resulted in a final serum dilution of 1:40 to 1: 29160. Next, the Vero E6 cells were washed once with PBS and the antibody-virus complexes were transferred to cells and incubated for 1 h at 37°C. Each serum sample and dilution was tested in duplicate and each assay included positive (pooled convalescent sera from BRBV infected mice), negative serum, and no serum controls. After 1 h at 37°C, the cells were washed once with PBS and 100 µL of MEM containing 2% FBS and 1% methylcellulose was added to each well. After 24 h at 37°C, the cells were fixed by adding 100 µL of 10% Formalin on top of the overlay (final concentration 5% Formalin) for 1 h at RT. Subsequently, cells were washed with PBS and the assay was developed as described above for the focus forming assay. The percent inhibition of BRBV infection was calculated by dividing the number of foci in the test sample by the average number of foci in the no serum control wells.

### Statistical analysis

Data were analyzed by using GraphPad Prism version 9.3.1 software. EC50 and IC50 values were calculated by using log (inhibitor) vs response in variable slope (four parameters) fixing both bottom and top constrain at 0 and 100 respectively.

## RESULTS

### Construction of a replication-competent VSV expressing BRBV GP

Since authentic BRBV is a biosafety level 3 (BSL3) pathogen, we sought to develop a tool to study aspects of the BRBV lifecycle at reduced biocontainment. To do this, we replaced the glycoprotein gene in a molecular clone of eGFP-expressing VSV with the GP gene of BRBV (VSV-BRBV) (**Fig 1A**). VSV-BRBV was capable of robust eGFP expression in Vero-CCL81 and grew to high titers (>10^8^ FFU/mL) with similar plaque size to VSV containing its native glycoprotein (**Fig 1B, C**).

**Figure 1:**
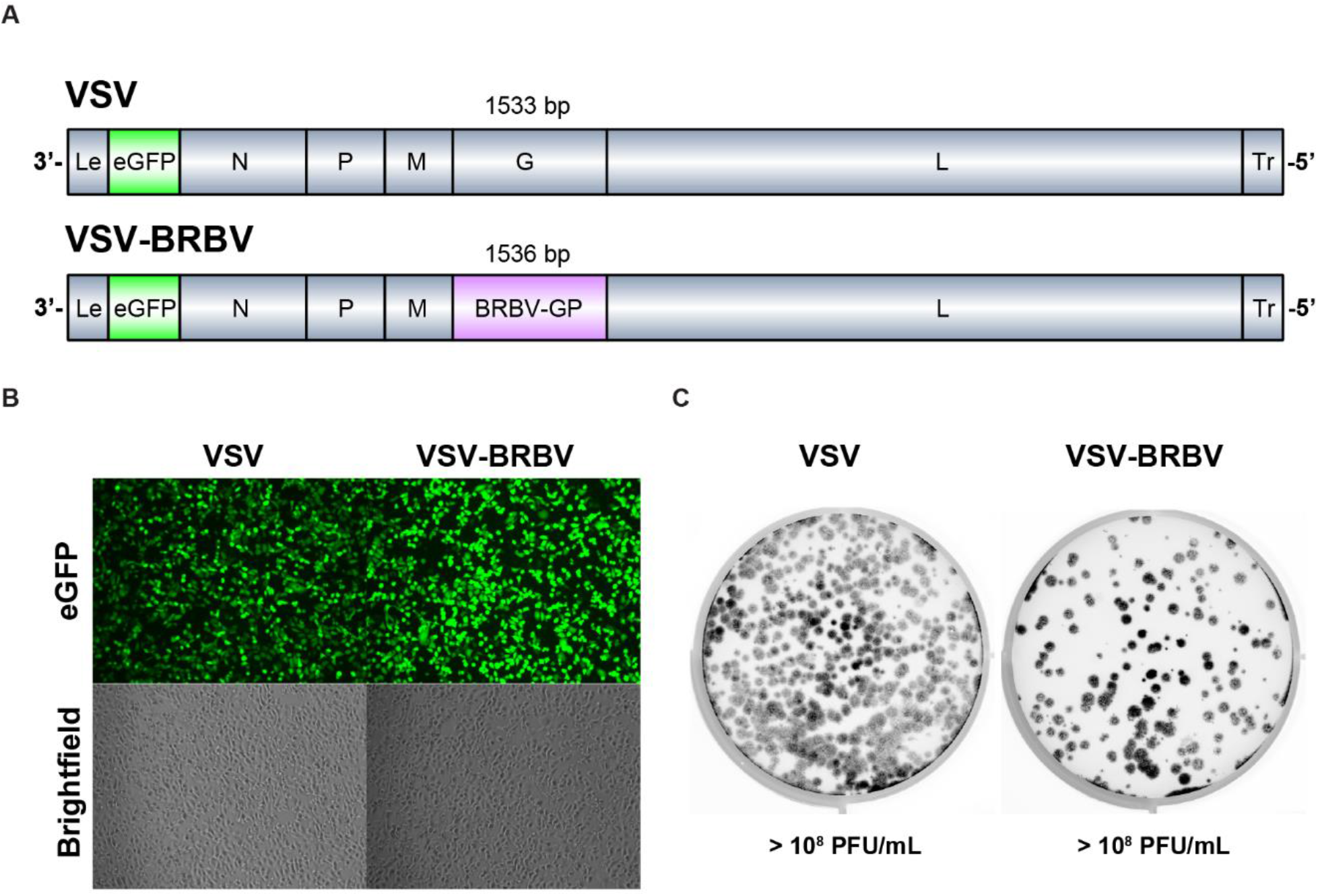
Generation of a replication-competent VSV expressing eGFP and BRBV GP. (**A**) Genome schematic. Genes encoded by each virus are to scale (except Le – Leader and Tr – Trailer) and are indicated by eGFP (enhanced green fluorescent protein), N (nucleoprotein), P (phosphoprotein), M (matrix protein), G or GP (glycoprotein), and L (large protein or polymerase). Glycoprotein gene length excludes the stop codon. (**B**) Vero-CCL81 were infected with VSV or VSV-BRBV at an MOI of 1 images were acquired 6 hours post infection (hpi). (**C**) Representative plaque assay of VSV or VSV-BRBV on Vero-CCL81 cells. Images were acquired 24 hpi.

### Development of humanized monoclonal antibodies against GP of BRBV

Eleven- to-twelve weeks-old C57BL/6 mice were immunized intramuscularly with inactivated BRBV followed by a single intramuscular boost with rGP of BRBV 33 days later. Sera were collected 21 and 5 days after the first and second immunization respectively (**Fig 2A**) and used to measure GP specific antibody responses by ELISA. Compared to the primary immunization, mice boosted with rGP of BRBV showed ∼6-fold increase in serum antibody titer for binding to BRBV rGP (**Fig 2B**). Five days after the boost, the spleens from one immunized and a control animal were isolated and processed for detection and isolation of BRBV GP-specific B cells. Nearly 2% of the ASC B-cells (CD4^-^, CD19^+^, IgD^lo^, CD95^+^, GL7^-^, CD138^+^) were specific for BRBV-GP (**Fig 2C**). No BRBV-GP specific ASC B-cells were detected in the naïve control animal. The BRBV-GP labelled B cells were then single cell sorted by FACS into 96-well plates containing lysis buffer. Amplification of VH, Vκ, and Vλ genes from a single B-cell sorted 96-well plate showed ∼70% productive V-domains (n= 66/94) which were later grouped into nine different clonal groups based the identity of their V and J alleles and the CDR3 region. One representative VH and VL gene from each clonal group were cloned into human IgG1 expression vectors, transfected in HEK293 cells, and purified. These mAbs are further referred to as BRBV-GP-A05, -A08, -A10, -B02, -B03, -B04, -E02, -E04, and -F07. Purified mAbs were tested for specificity and affinity to rGP by ELISA (**Fig 2D**). All nine antibodies were specific for BRBV-GP, three (BRBV-GP-A05, -A08, and -B02) of which demonstrated binding affinity to BRBV rGP with an EC50 < 500 ng/mL.

**Figure 2:**
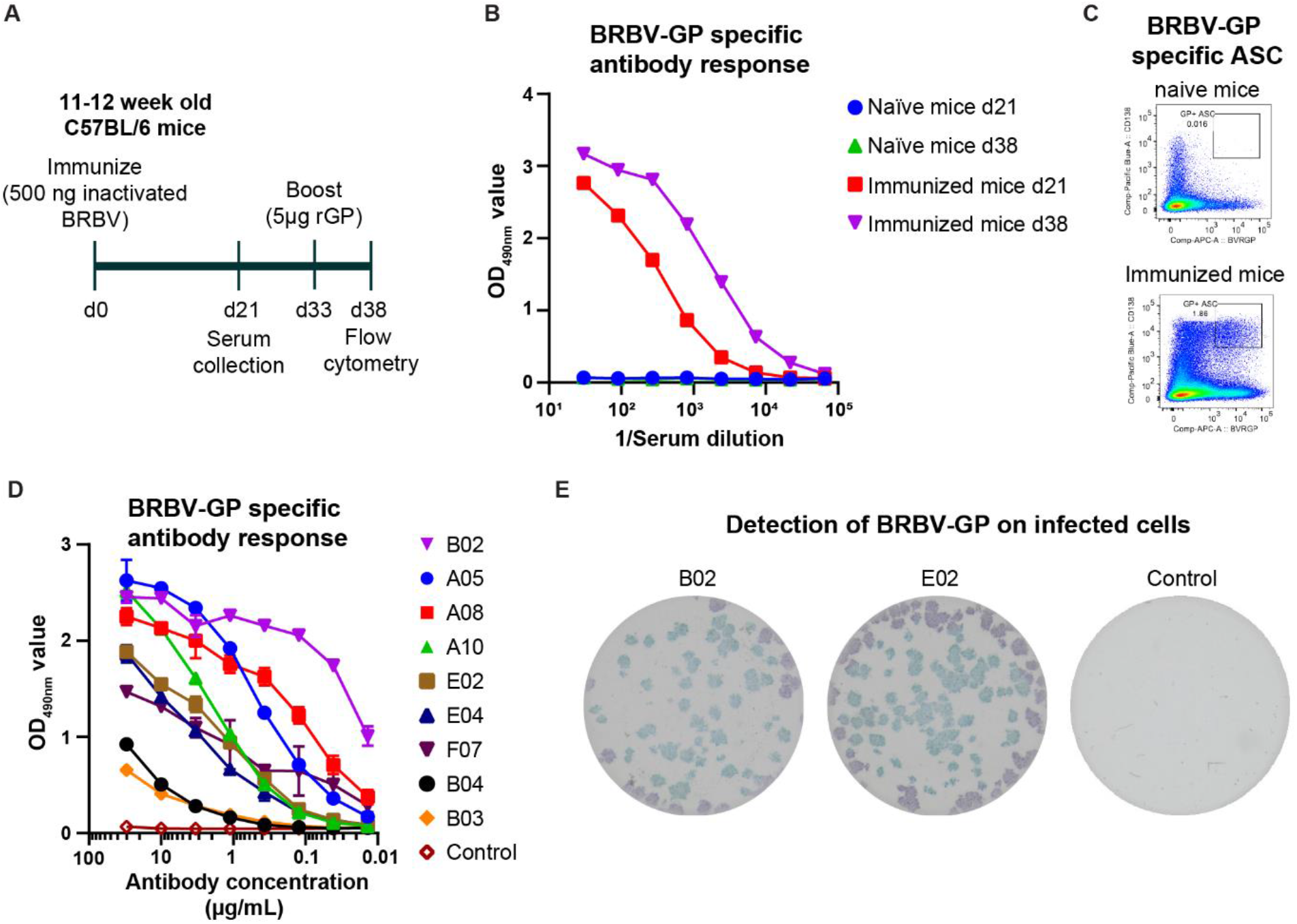
Generation and characterization of humanized monoclonal antibodies against BRBV GP. (**A**) Immunization regimen. 11-12 weeks old C57BL/6 mice were immunized intramuscularly with 500 ng BPL-inactivated BRBV, followed by boosting with 5 µg of rGP of BRBV 33 day later. Sera was collected at days 21 and 38. Spleens were harvested five days post boost (day 38) (**B**) IgG serum Ab ELISA for rGP of BRBV for naïve and immunized mice at days 21 and 38. Results are average of two independent experiments. (**C**) Representative plots of BRBV-GP specific ASC B-cells (CD4^-^, CD19^+^, IgD^lo^, CD95^+^, GL7^-^, CD138^+^, BRBV-GP^+^ live singlet lymphocytes) in naïve mice (top) and BRBV-immunized mice (bottom) (**D**) ELISA measuring binding of nine mAbs to rGP of BRBV. Each line is a different antibody and the results are the average of two independent experiments. (**E**) Binding of mAb B02 and E02 to BRBV-infected cells. Vero cells were infected with ∼100 infectious units of BRBV for 24 h, fixed with 5% formalin, and virus-infected cells were visualized by incubating the cells with 2-3 µg/mL of mAbs B02 and E02, or a control. Images are representative of 2 experiments.

The mAbs were further evaluated in a FFA with BRBV for their ability to detect surface expressed GP on infected cells. In cells infected with BRBV, only two of the nine tested mAbs (B02, and E02) were able to detect the GP protein on the surface of infected cells and produce distinct foci in this assay (**Fig 2E**). The remaining seven mAbs were unable to detect GP expression in BRBV-infected Vero E6 cells. Of the two mAbs, E02 produced the most distinct and intense foci in the BRBV FFA (**Fig 2E**). This data combined with the ELISA data (**Fig 2D**), demonstrate that we produced the first BRBV specific monoclonal antibodies that can be used for diagnostic assays.

### A rapid, eGFP-based neutralization assay for BRBV

To facilitate throughput and reduce the costs associated with BSL3 work, we developed a rapid and high-throughput eGFP-based neutralization assay using a replication competent BRBV-GP pseudotyped VSV that also expressed eGFP (VSV-BRBV). To demonstrate the utility of this virus, cells were inoculated with 200 FFU of VSV-BRBV with or without sera containing neutralizing antibodies against BRBV. After 8 h without serum, eGFP expression is clearly visible in a subset of the cells (**Fig 3A, left panel**). The addition of mouse sera obtained from naïve C57BL/6 animals did not inhibit the infection and replication of VSV-BRBV and similar number of eGFP positive cells were visible (**Fig 3A, middle panel**). In contrast, the addition of serum collected 22 days after inoculation of C57BL/6 mice with 10^4^ infectious units of BRBV, completely neutralized VSV-BRBV and no eGFP-positive cells were detected (**Fig 3A, right panel**). Analysis of the positive and negative control sera show a significant difference (*P* < 0.0001) in neutralization of VSV-BRBV (**Fig 3B**). Combined, these data show that VSV-BRBV can be used as a high-throughput alternative for live BRBV to detect BRBV neutralizing antibodies.

**Figure 3:**
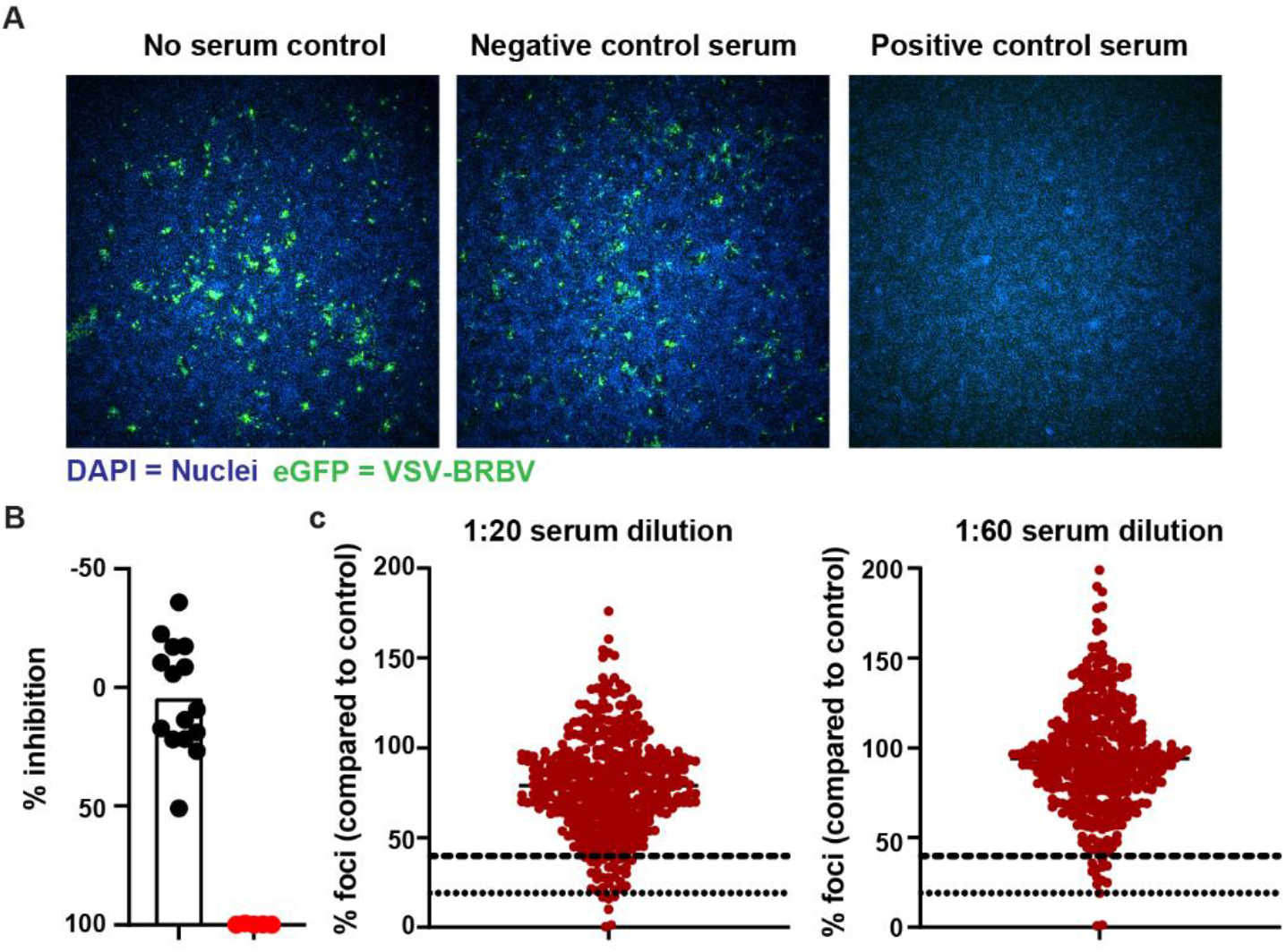
Development of a rapid eGFP-based BRBV neutralizing assay. (**A**) Effective neutralization of VSV-BRBV by BRBV-neutralizing antibodies. Monolayers of Vero cells incubated for 8 h with VSV-BRBV, expressing eGFP, in the presence of mouse serum-containing BRBV neutralizing antibodies (positive control), control mouse serum, or no serum were visualized with DAPI (blue color). VSV-BRBV infected cells were visualized by the expression of GFP from the VSV genome. Representative images of the no serum (left), negative serum (middle), and positive serum (right) controls are shown at 4x magnification. (**B**) Normalized VSV-BRBV neutralization (% inhibition compared to no serum control) of the positive and negative control sera. Each symbol is an individual mouse serum sample (n = 15 negative control, n = 5 positive control) (**C**) Normalized VSV-BRBV infection (% inhibition compared to no serum control) of 440 human serum samples at a 1:20 (left panel) and 1:60 (right panel) dilution of the sera. Each symbol is a serum sample. The dotted lines represent the cut-offs for 60% and 80% inhibition.

A total of 440 human serum samples were tested in the rapid VSV-BRBV based neutralization assay at two different dilutions of the sera (1:20 and 1:60). The inhibition of VSV-BRBV infection was normalized to the no serum control wells included in each assay. The vast majority of the sera did not neutralize VSV-BRBV at either the 1:20 or 1:60 dilution of the sera (**Fig 3C**). Interestingly, three human serum samples demonstrated > 80% reduction in VSV-BRBV infection at both serum dilutions (**Fig 3C**). In addition, 54 and 14 sera were identified as potentially positive (> 60%, but < 80% inhibition) at the 1:20 and 1:60 dilution of the sera respectively (**Fig 3C**). The 14 samples, identified as potentially positive in 1:60 dilution, were also potentially positive in 1:20 dilution and selected for further testing in the BRBV FRNT assay together with 3 positive samples.

### Neutralization of BRBV by human serum samples

To confirm our findings and demonstrate neutralizing of live BRBV by antibodies in human sera, we developed a FRNT for BRBV using the BRBV-GP specific mAb (E02) described above. Sera collected from naïve mice was unable to neutralize BRBV and the number of foci was comparable to the no serum controls (**Fig 4A, left panel**). In contrast, convalescent sera from mice infected with BRBV was able to potently neutralize live BRBV (**Fig 4A, right panel**). The IC50 for these sera ranged between 1:500 and 1:1350.

**Figure 4:**
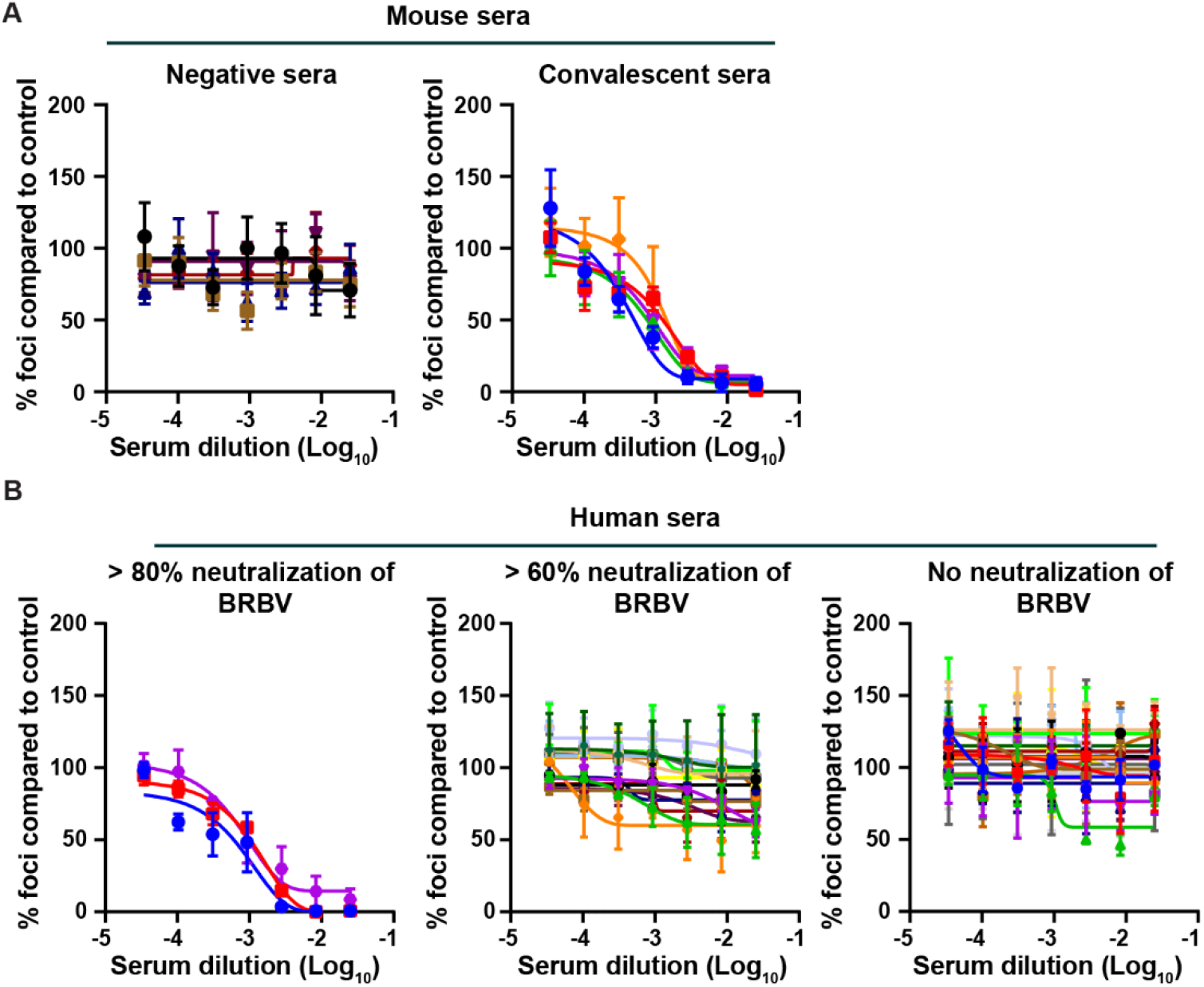
Detection of serum neutralizing antibodies against BRBV in human serum samples. A focus reduction neutralization test (FRNT) was developed for BRBV. Serially diluted heat-inactivated human or mouse sera (starting at 1:40 dilution) was incubated for 1 h with 100 focus forming units of BRBV prior to adding it to Vero cells. One h later, the inoculum was removed, the cells were overlaid with 1% methylcellulose, and incubated for 24 h. Clusters of infected cells (foci) were visualized using the E02 mAb as described in the Materials and Methods section. The number of foci are normalized to no serum controls. (**A**) *Left*. FRNT with serum from mock-infected control animals. No BRBV neutralizing activity was detected in the negative control sera. *Right*. FRNT demonstrating virus-neutralizing activity in convalescent serum collected from C57BL/6 mice 22 days after inoculation with 10^4^ PFU of BRBV. Neutralization of BRBV was detected in all five mouse serum samples. (**B**) FRNT on human serum samples with distinct neutralizing activities in the rapid assay. Human serum samples showing complete neutralization (0-20%, *left*), partial neutralization (20-40%, middle), or no neutralization (100%, right) in the VSV-BRBV rapid assay were tested by FRNT. Three human sera were able to neutralized BRBV by FRNT.

Next, the BRBV FRNT assay was used to test the 17 human serum samples that previously demonstrated complete or partial neutralization of VSV-BRBV (**Fig 3C**). The three serum samples that strongly inhibited (> 80% inhibition) VSV-BRBV also inhibited BRBV in the FRNT assay (**Fig 4B, left panel)**. The IC50 values for these three serum samples were 1:2631, 1:1259 and 1:938. The remaining 14 human serum samples that partially inhibited VSV-BRBV infection (> 60%, but <80% inhibition) did not neutralize BRBV in the FRNT assay (**Fig 4B, middle panel**). As a control we tested 19 human serum samples that did not neutralize VSV-BRBV. None of the 19 control sera were able to inhibit BRBV infection and their IC50 values were at the limit of detection (1:40) (**Fig 4B, right panel)**. Collectively, these data demonstrate that BRBV infections occur in the St Louis community and that the seroprevalence of BRBV neutralizing antibodies is ∼0.7%.

## DISCUSSION

Here we defined the first seroprevalence of BRBV serum neutralizing antibodies in human sera collected from the greater St. Louis Metropolitan area. We developed two virus neutralization assays, a rapid VSV-BRBV eGFP based neutralization assay, and a BRBV focus reduction neutralization assay, to systematically evaluate the presence of BRBV neutralizing antibodies in human serum. These assays identified 3 (0.7%) human sera that contained BRBV specific neutralizing antibodies. These finding indicate that the incidence of BRBV infection in the St. Louis region in Missouri, US is higher than previously expected based simply on confirmed cases that have been reported.

BRBV is classified as a BSL3 pathogen and needs specialized laboratories, training, and equipment to work with. As such, limited labs in the US and around the world have the capacity to work with live BRBV. To facilitate the testing of BRBV-GP specific countermeasures and determine the seroprevalence of neutralizing antibodies in humans and wildlife, we developed a rapid eGFP-based neutralization assay using replication-competent VSV expressing both eGFP and BRBV GP. This virus can be used at lower biosafety containment levels (BSL2). Importantly, compared to previously described methods such as PRNT, both the eGFP-based rapid assay and the BRBV FRNT can be used to evaluate BRBV neutralizing antibodies within three days. Thus the turnaround time is greatly improved.

One of the major limitation of studying emerging pathogens is the lack of reagents such as mAbs, recombinant proteins, reporter viruses etc. This is the first report describing mAbs specific for GP of BRBV. Two of these mAbs were able to detect GP expression on cells infected with BRBV. Unfortunately, these antibodies did not neutralize BRBV, suggesting they do not bind the receptor binding site of the virus [3].

BRBV and HRTV are classified as emerging infectious diseases that are transmitted via ticks in the US. A previous study demonstrated a 0.9% seroprevalence of HRTV specific neutralizing antibodies in northwestern Missouri, US, where human cases and infected ticks have been identified [23]. Our study demonstrates a 0.7% seroprevalence of BRBV specific neutralizing antibodies in individuals in the St. Louis area. This is the first time that BRBV neutralizing antibodies have been reported in individuals without a prior history of BRBV infection. These data also suggest that the true infection burden of BRBV in humans is much higher than previously expected. The seroprevalence in this cohort may underestimate the true infection rate and burden. First, our human serum cohort is from the greater St. Louis area and is likely composed primarily of urban populations that are less likely to be exposed to BRBV infected ticks. Future seroprevalence studies with sera from individuals living in rural areas where BRBV infection in ticks have been reported are needed. Second, the presence of detectable levels of neutralizing antibodies is a relatively high threshold for determining prior infection with any virus including BRBV. For example, there are many individuals previously infected with the influenza virus or severe acute respiratory syndrome coronavirus 2 (SARS-CoV-2) who have no detectable serum neutralizing antibodies. Moreover, the true infection burden of BRBV in US could be underestimated due to lack of diagnostic testing, unknown virus geographical distribution, and unknown rate of asymptomatic infections. Combined with the widespread distribution of the lone star tick and the reported presence of BRBV-specific antibodies in North Carolina, we expect the true infection burden of BRBV to be much higher than what was previously expected based on the number of confirmed BRBV infections per year.

Three human sera showed the highest neutralization activity (> 80% inhibition) in the VSV-BRBV eGFP-based assay. These same samples were confirmed positive for BRBV-specific neutralizing antibody by FRNT. These findings suggest that the rapid assay and the BRBV FRNT are consistent in detecting the true seropositive samples and that our cut-off of 80% is rigorous. These two BRBV neutralization assays will expand our knowledge of BRBV seroprevalence in humans and in wild animals. Previous studies have shown that serum from white-tailed deer, raccoons, domestic dogs, horses, and eastern cottontails also contains BRBV neutralizing antibodies [12, 13]. The assays developed in this study will facilitate additional testing of animal species in BRBV endemic and non-endemic areas and help identify the breadth of host species affected by this virus.

In conclusion, our study established two neutralization assays for BRBV and for the first time evaluated BRBV seroprevalence in a cohort of human sera. We found 3 people that were positive for BRBV neutralizing antibodies.

## Data Availability

All data produced in the present work are contained in the manuscript

## ACKNOWLEDGEMENTS

We thank Maxene Ilagan at Washington University School of Medicine High Throughput Center for supporting with imaging. This study was supported by NIH/NIAID grants R21 AI151170 and U01 AI151810. P.W.R. is supported by 1F31AI154710-01A1.

## AUTHOR CONTRIBUTIONS

G.B. and P.W.R. designed experiments. G.B. performed the neutralization assays. H.H. and A.E. generated and tested the monoclonal antibodies. P.W.R. and S.P.J.W. generated and characterized the VSV-BRBV virus. Y.D. and D.F. generated the recombinant glycoproteins. D.W. collected human serum samples. A.C.M.B. obtained the funding. G.B., H.H., P.W.R., and A.C.M.B. wrote the manuscript, and all authors edited the final version.

